# Increased Risk of Portal Hypertension-Related Complications in Those with History of Bariatric Surgery and Alcohol-Associated Hepatitis

**DOI:** 10.64898/2026.03.12.26348268

**Authors:** Brandon Havranek, Thomas Z. Rohan, Chenab K. Khakh, Rachel Redfield, Dina Halegoua-DeMarzio

## Abstract

**Background and Objectives:** Bariatric surgery is a highly effective obesity treatment, yet it may predispose individuals to alcohol-related liver injury. While altered ethanol metabolism following procedures like Roux-en-Y gastric bypass (RYGB) is well described, the long-term hepatic consequences, particularly the risk of portal hypertension in patients who develop alcohol-related hepatitis (AH,) remain poorly defined.

**Methods:** Using the TriNetX US Collaborative Network, we identified adult patients diagnosed with AH or alcohol-related cirrhosis. We compared outcomes between patients with a history of RYGB or sleeve gastrectomy (SG) who subsequently developed AH (Bariatric+AH group) and those with AH and no history of bariatric surgery (AH-only group). Propensity score matching was performed on over 44 demographic, clinical, and laboratory variables. Cox proportional hazards models and Kaplan-Meier survival curves were used to estimate the risk of clinically significant portal hypertension (PH) events, liver transplantation, and all-cause mortality at three-, five-, and seven-year follow-ups.

**Results:** After matching, 772 patients were included in each cohort. At 7 years post-index event, the Bariatric + AH group exhibited a significantly higher risk of PH-related complications compared to the AH-only group (HR 1.519; 95% CI, 1.15–2.005; p = 0.003). No significant differences were observed in liver transplantation (HR 1.412; 95% CI, 0.850–2.346; p = 0.181) or all-cause mortality (HR 1.085; 95% CI, 0.904–1.303; p = 0.381). These findings were consistent across all follow-up intervals.

**Conclusion:** Bariatric surgery is associated with an increased long-term risk of portal hypertension in patients who develop alcohol-related hepatitis despite similar mortality and transplantation rates. These findings underscore the need for targeted postoperative counseling, liver-focused surveillance strategies, and integration of hepatologic risk assessment into metabolic surgery care pathways.

## Introduction

Obesity rates in the United States continue to climb, with an estimated prevalence of over 41.9% among adults, prompting a dramatic rise in the popularity of weight loss interventions, particularly bariatric surgery.^1^ Bariatric surgery is one of the most effective interventions for weight loss, demonstrating long-term improvements in metabolic health, cardiovascular outcomes, and overall survival.^2^ However, it is well established that bariatric surgeries can alter gastrointestinal anatomy and physiology in ways that impair the absorption and metabolism of certain nutrients and substances, including ethanol.^3^

Prior studies have shown that blood ethanol levels peak higher, faster, and take longer to return to baseline after bariatric surgery, particularly following Roux-en-Y gastric bypass (RYGB). This phenomenon can be attributed to bypassing alcohol dehydrogenase-rich regions of the stomach and small intestine.^4,5^ This altered pharmacokinetic profile increases susceptibility to rapid intoxication. The risk appears less defined for other procedures, such as sleeve gastrectomy (SG), though some evidence suggests a mild elevation in alcohol misuse risk.^6^

The potentially increased risk of adverse health outcomes due to alcohol, coupled with the psychological and behavioral vulnerabilities common in the postoperative population, makes evaluating alcohol-mediated liver complications after bariatric surgery a critical research priority. While some studies have linked bariatric surgery to increased risk of alcohol-related cirrhosis, hepatitis, and alcohol use disorder (AUD), particularly in women and younger individuals, most have lacked sufficient follow-up time to assess long-term liver-related outcomes such as clinically significant portal hypertension or need for liver transplantation.^6,7^ Additionally, patients with a history of bariatric surgery may be at risk of presenting with more severe forms of alcohol-associated liver disease (AALD), such as alcohol-related hepatitis (AH). Yet, the outcomes of these patients, specifically with regard to mortality, transplantation, and hepatic decompensation remain poorly characterized.

The primary objective of this study is to investigate whether individuals who have undergone bariatric surgery and subsequently developed alcohol-related hepatitis (AH) experience worse clinical outcomes compared to those with AH but no history of bariatric surgery. Given the known alterations in alcohol metabolism following procedures such as Roux-en-Y gastric bypass (RYGB) and the potential for increased susceptibility to alcohol-related liver damage, this study aims to evaluate whether these physiological changes translate into greater clinical severity. Specifically, the study will assess the incidence of clinically significant portal hypertension (PH) events, liver transplantation, and all-cause mortality among patients with AH, stratified by bariatric surgery status. By examining these outcomes, the research seeks to clarify the long-term hepatic consequences of bariatric procedures in the context of alcohol-related liver disease and provide insights that could inform risk stratification, preoperative counseling, and long-term monitoring of post-bariatric patients.

## Methods

### Study Population

All adult patients (aged > 18 years) with a history of sleeve gastrectomy (SG) or Roux-en-Y gastric bypass (RYGB) and diagnosis of alcoholic hepatitis or alcoholic cirrhosis were queried in the TriNetX US Collaborative Network which contains over 60 US health care organizations. Patients who met the inclusion criteria were divided into two cohorts. The study group cohort contained patients who underwent SG or RYGB and subsequently developed alcohol-related hepatitis (AH) or alcohol-related cirrhosis following surgery. The second control cohort (AH only) included patients with AH or alcohol-related cirrhosis and no prior history of RYGB or SG. ICD-10-CM and CPT codes used for the cohort criteria are supplied in the supporting information.

### Propensity Score Matching and Covariates

To ensure the groups were comparable, 1:1 propensity score matching (PSM) was conducted, incorporating over 44 variables such as age, demographics, comorbid conditions, laboratory parameters, and medication usage (a complete list of variables included in the matching process is provided in the supplementary material). Following the matching process, each cohort consisted of 772 patients. Propensity scores for individuals in both cohorts were calculated by performing logistic regression on the respective datasets within the TriNetX platform. This regression analysis was executed using Python version 3.6.5 (Python Software Foundation), utilizing the NumPy and sklearn libraries, and independently validated using R version 3.4.4 (R Project for Statistical Computing) to maintain reproducibility. Matching was performed using a greedy nearest-neighbor algorithm, applying a caliper width of 0.1 pooled standard deviations. Prior to matching, the order of the covariate matrices was randomized to mitigate potential bias related to row sequencing. Variables included in the propensity score model were selected to adjust for confounders measured up to one day before the index event.

### Study Outcomes

Our primary objective was to assess the incidence of clinically significant portal hypertension (PH) events defined as the first occurance of portal hypertension, esophageal varices (with and without bleeding), gastric varices, ascites, hepatorenal syndrome, hepatic failure, jaundice, chronic passive congestion of liver, and hepatic encephalopathy. The secondary outcomes included incidence of all-cause mortality and liver transplantation. These study outcomes were analyzed at 3-, 5-, and 7-year intervals following the index event. Patients were excluded from the analysis if they had experienced any defined outcomes before the index date. ICD-10-CM and CPT codes used to identify study outcomes are supplied in the supporting information.

### Statistical Analysis

All statistical analyses were performed in real time through the TriNetX analytics platform. Continuous variables were summarized using means and standard deviations (SD), whereas categorical variables were described as counts and percentages. Propensity score matching (PSM) was applied to create comparable groups, and the balance of covariates post-matching was evaluated using standardized mean differences (SMD), with a predefined threshold of 0.10 to indicate acceptable balance. SMDs were selected over p-values as they are less influenced by sample size and more appropriate for assessing covariate balance.^8^ Cox proportional hazards regression was utilized to calculate hazard ratios (HRs) and 95% confidence intervals (CIs) for the outcomes of interest.^9^ The proportional hazards assumption was assessed, and all Cox analyses were conducted using the survival package in R version 3.2.3. Adjusted HRs and 95% CIs, controlling for baseline characteristics, were reported for each analysis. The results were cross-validated using SAS software version 9.4 (SAS Institute) to ensure consistency. Kaplan-Meier survival curves were generated to estimate outcome-free survival probabilities at 3, 5, and 7 years following the index event. Patients were censored at the end of the observation period or on the day following their last documented encounter. P-values associated with Kaplan-Meier curves were calculated using the log-rank test, with statistical significance defined as a two-sided alpha level below 0.05. Data analysis was completed in November 2023.

### Ethics Statement

All study procedures adhered to applicable ethical guidelines and regulatory requirements. The analysis was conducted using data obtained solely from the TriNetX Research Network, which aggregates de-identified patient information from participating healthcare institutions. These institutions have secured the necessary permissions, consents, and legal authority to contribute data to TriNetX under Business Associate Agreements, ensuring anonymity and restricting the data’s use to research purposes only. This framework guarantees that no individual patients can be identified, either directly or indirectly, thereby maintaining compliance with the Health Insurance Portability and Accountability Act (HIPAA) privacy standards. Based on these safeguards and the retrospective nature of the study, the Thomas Jefferson University Institutional Review Board (IRB) determined that formal informed consent was not required. No identifiable data pertaining to patients or contributing healthcare entities were accessed or disclosed in the conduct of this study.

## Results

### Patient Charactertistics

A total of 789 patients with history of SG or RYGB who went on to developed AH were identified, while 289,022 patients with solely AH and no history of SG or RYGB were identified. After PSM to account for differences between the two study groups (see methods), a total of 772 patients were included in each group. In the bariatric + AH group at index the mean (SD) age was 53.8 (12.0), 399 (51.7%) female, 510 (66.1%) white, while in the control group (AH only) the mean (SD) age was 54.0 (13.0), 409 (53.0%) female, and 516 (66.8%) white. Both groups were well-matched after PSM (majority SMD < 0.10) (Table 1). The only imbalances remaining after PSM (SMD > 0.10) included the use of antiarrhythmics, antilipemic agents, calcium channel blockers, and other hypertensive agents (Table 1).

**Table 1.** Baseline characteristics of bariatric + AH group and AH only group after propensity score matching.

| Patients, No. (% of cohort) |  |  |  |
| --- | --- | --- | --- |
| Characteristics | bariatric + AH (n = 772) | AH only (n = 772) | Standard mean difference |
| **Age, mean (SD)** | 53.8 (12.0) | 54.0 (13.0) | 0.011 |
| **Sex** |  |  |  |
| Female | 399 (51.7) | 409 (53.0) | 0.026 |
| Male | 350 (45.3) | 341 (44.2) | 0.023 |
| **Race and ethnicity** |  |  |  |
| Non-Hispanic White | 510 (66.1) | 516 (66.8) | 0.016 |
| Non-Hispanic Black | 106 (13.7) | 97 (12.6) | 0.035 |
| Hispanic or Latino | 61 (7.9) | 75 (9.7) | 0.006 |
| **BMI, mean (SD)** | 30.0 (8.2) | 30.5 (7.6) | 0.065 |
| **Nicotine dependence** | 299 (38.7) | 299 (38.7) | 0.001 |
| **Comorbidities** |  |  |  |
| Type 2 diabetes | 286 (37.0) | 281 (36.4) | 0.013 |
| Hypertensive diseases | 555 (71.9) | 552 (71.5) | 0.009 |
| Hyperlipidemia | 286 (37.0) | 271 (35.1) | 0.040 |
| Atrial fibrillation and flutter | 80 (10.4) | 105 (13.6) | 0.100 |
| Abnormalities of heart beat | 321 (41.6) | 306 (39.6) | 0.040 |
| Chronic lower respiratory disease | 279 (36.1) | 282 (36.5) | 0.008 |
| Cerebrovascular diseases | 91 (11.8) | 88 (11.4) | 0.012 |
| Ischemic heart diseases | 181 (23.4) | 200 (25.9) | 0.057 |
| Other forms of heart disease | 390 (50.5) | 387 (50.1) | 0.008 |
| Diseases of veins, lymphatic vessels and lymph nodes, not elsewhere classified | 265 (34.3) | 269 (34.8) | 0.011 |
| Diseases of arteries, arterioles, and capillaries | 151 (19.6) | 163 (21.1) | 0.039 |
| Other diseases of the respiratory system | 282 (36.5) | 289 (37.4) | 0.019 |
| **Blood pressure, mean (SD), mmHg** |  |  |  |
| Systolic | 123.1 (22.2) | 124.5 (22.1) | 0.064 |
| Diastolic | 73.7 (15.0) | 72.8 (15.2) | 0.065 |
| **HbA<sub>1c</sub>, % (SD)** | 5.8 (1.6) | 6.0 (1.6) | 0.019 |
| **Liver function panel, mean (SD)** |  |  |  |
| ALT, U/L | 48.1 (55.0) | 50.5 (167.6) | 0.019 |
| AST, U/L | 81.7 (97.1) | 88.5 (428.9) | 0.022 |
| ALP, U/L | 153.4 (151.5) | 136.4 (94.2) | 0.001 |
| Total bilirubin, mg/dL | 2.0 (3.7) | 1.8 (3.3) | 0.042 |
| Albumin, g/dL | 3.3 (0.8) | 3.4 (0.8) | 0.073 |
| **Kidney function, mean (SD)** |  |  |  |
| Creatinine, mg/dL | 0.9 (1.0) | 1.1 (1.3) | 0.052 |
| **Lipid panel, mean (SD)** |  |  |  |
| Cholesterol, mg/dL | 161.5 (53.7) | 162.4 (55.1) | 0.016 |
| Triglyceride, mg/dL | 127.3 (98.0) | 147.1 (131.6) | 0.017 |
| LDL, mg/dL | 85.0 (43.1) | 87.2 (40.4) | 0.016 |
| **Coagulation profile, mean (SD)** |  |  |  |
| PT, s | 14.7 (6.5) | 14.8 (5.1) | 0.012 |
| **Medications** |  |  |  |
| Antiarrhythmics | 628 (81.3) | 558 (72.3) | 0.216 |
| Antilipemic agents | 259 (33.5) | 311 (40.3) | 0.140 |
| β-Blockers | 488 (63.2) | 462 (59.8) | 0.069 |
| ACE inhibitors | 263 (34.1) | 272 (35.2) | 0.025 |
| Angiotensin II inhibitors | 142 (18.4) | 151 (19.6) | 0.030 |
| Calcium channel blockers | 247 (32.0) | 305 (39.5) | 0.157 |
| Antihypertensives, other | 308 (39.9) | 262 (33.9) | 0.124 |
| Antianginals | 141 (18.3) | 166 (21.5) | 0.081 |
| Diuretics | 497 (64.4) | 499 (64.6) | 0.005 |
| Oral hypoglycemic agents | 177 (22.9) | 194 (25.1) | 0.052 |
| Insulin | 329 (42.6) | 312 (40.4) | 0.045 |
| **Supplements** |  |  |  |
| Vitamin D | 368 (47.7) | 375 (48.6) | 0.018 |
| Vitamin E | 77 (10.0) | 79 (10.2) | 0.009 |

### Primary Outcome

Patients in the bariatric + AH group were compared to their matched control AH only group using a Cox proportional hazards model to evaluate the risk of new onset PH events. The patients in the bariatric + AH group demonstrated a significantly higher risk of experiencing clinically significant PH events at 3 years (HR, 1.533; 95% Cl, 1.147-2.048), 5 years (HR, 1.532; 95% Cl, 1.157-2.03), and 7 years (HR, 1.519; 95% Cl, 1.15-2.055) after the index event (Figure 1).

**Figure 1.**
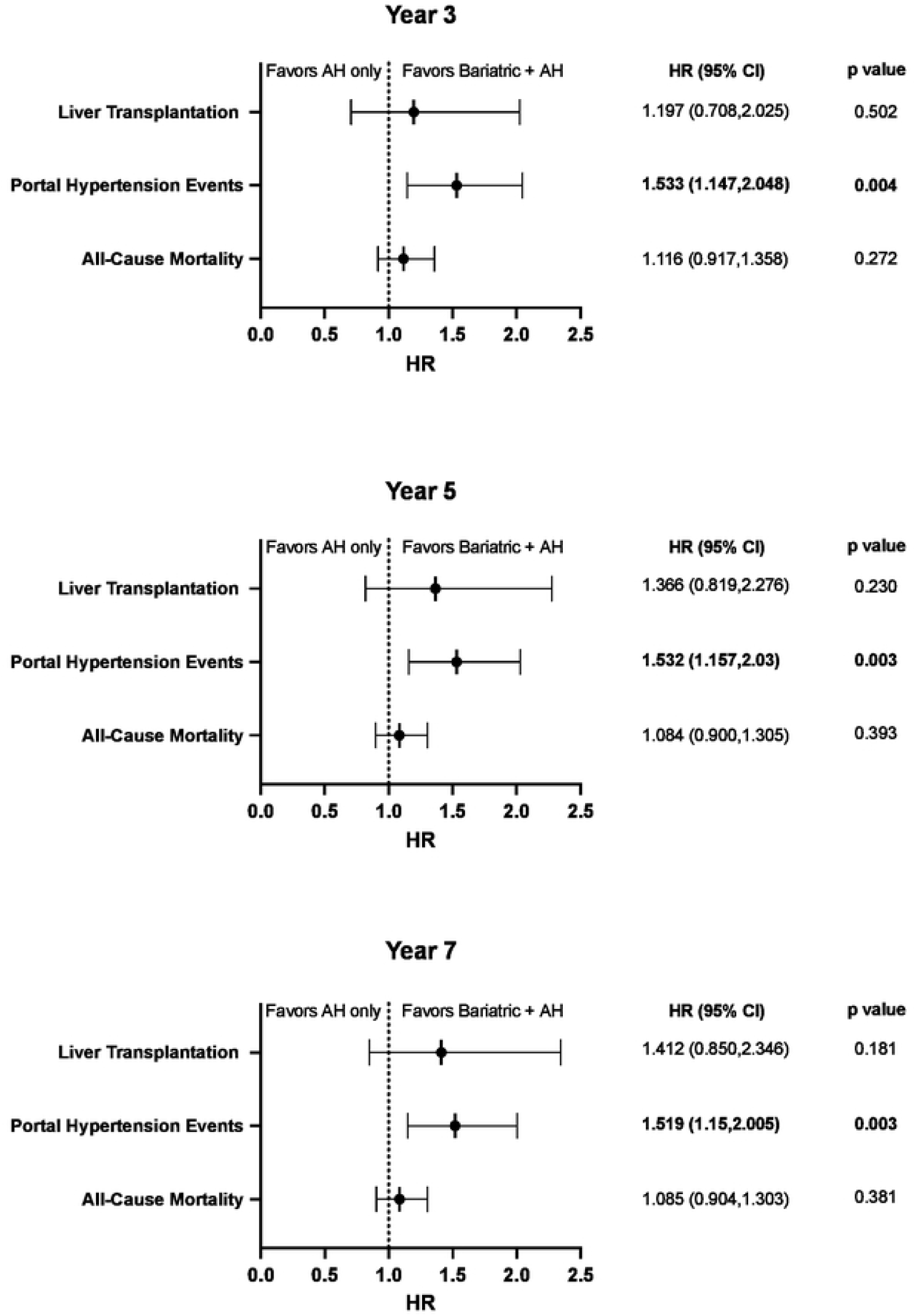
Association of patients with bariatric surgery and alcohol-associated hepatitis with liver transplantation, clinically significant portal hypertension events, and all-cause mortality.^a^ ^a^p values were calculated from Kaplan-Meier survival curves and were conducted using the log-rank test.

### Secondary Outcomes

Additonally, we compared patients in the bariatric + AH group to their matched control AH only group in regards to liver transplantation. There were no statistically significant differences noted between the bariatric + AH group vs AH only control group in regards to liver transplantation at 3 years (HR, 1.197; 95% Cl, 0.708-2.025), 5 years (HR, 1.366; 95% Cl, 0.819-2.276), and 7 years (HR, 1.412; 95% Cl, 0.850-2.346) post index date (Figure 1). The same analysis was performed analyzing the outcome of all-cause mortality. Again, no significant differences were observed when comparing the two groups at 3 years (HR, 1.116; 95% Cl, 0.917-1.358), 5 years (HR, 1.084; 95% Cl, 0.900-1.305), and 7 years (HR, 1.085; 95% Cl, 0.904-1.303) after the index event (Figure 1).

## Discussion

In our analysis of a large patient population, we found that bariatric surgery patients who subsequently developed alcohol-related hepatitis were at an elevated risk of sequelae of portal hypertension compared to those with alcohol-related hepatitis and no history of bariatric surgery. There was no observed difference in rates of liver transplantation or all-cause mortality. These findings were consistent across three-, five-, and seven-year intervals, suggesting a constant risk trajectory. Portal hypertension is associated with substantial morbidity, including variceal bleeding, ascites, and hepatic encephalopathy. It can be a turning point in the progression of chronic liver disease overall, making it a vital area of research for patient outcomes.

Our findings reinforce a growing body of literature linking bariatric procedures to increased alcohol-related complications. Kim et al.^5^ identified RYGB as a significant risk factor for new alcohol-related liver diagnoses, while Mellinger et al.^6^ reported an increased incidence of alcohol-related cirrhosis and misuse, especially in women. Similarly, Anugwom et al.^7^ noted worse clinical outcomes in patients with RYGB hospitalized for alcohol-associated hepatitis. Our study adds to this knowledge by uniquely quantifying the longitudinal risk of portal hypertension in a well-matched US cohort, providing time-stratified estimates of clinically significant portal hypertension events over seven years.

The pathophysiological mechanisms underpinning these findings likely involve anatomical and hormonal changes following bariatric surgery. While procedures like RYGB reduce the volume of alcohol that can be consumed, they also alter ethanol metabolism by bypassing gastric alcohol dehydrogenase, leading to higher and more rapid peak blood alcohol concentrations.^4^ In addition, surgery-induced changes to the gut-liver axis, such as shifts in GLP-1, bile acid signaling, and gut permeability, that may further potentiate hepatic inflammation and injury in the setting of alcohol exposure.^10^ These factors collectively suggest a unique vulnerability of the post-bariatric population to progressive liver disease. With rising rates of both obesity and alcohol use disorders, the convergence of these two epidemics poses a serious and under-recognized threat to liver health.

By identifying a clear association between prior bariatric surgery and the future development of portal hypertension in patients with alcohol-related hepatitis, our study highlights the need for integrated, preventive strategies across surgical and hepatologic care.

There are some limitations worth noting. The retrospective design and reliance on ICD coding may limit sensitivity for outcome capture, and the absence of detailed alcohol use data restricts our ability to disentangle behavioral versus metabolic contributors to disease progression. Additionally, we could not assess post-surgical weight changes, nutritional status, or procedural complications, all of which may influence liver trajectory. Nevertheless, the robustness of our findings across matched cohorts and extended follow-up enhances their interpretability. Future work should explore the role of preoperative risk stratification for alcohol misuse, develop predictive models incorporating metabolic and psychosocial variables, and assess the potential impact of structured postoperative liver monitoring in high-risk individuals. From liver function testing to behavioral counseling, multidisciplinary care could become the new bariatric follow-up care standard.

## Conclusion

In conclusion, while bariatric surgery remains a powerful tool for combating obesity, its long-term hepatic consequences, especially in the context of alcohol use, demand careful attention. The elevated risk of portal hypertension-related complications observed in this study underscores the importance of counseling, follow-up, and interdisciplinary care strategies to mitigate long-term morbidity in this vulnerable population.

## Data Availability

All analyses were performed using the TriNetX Research Network platform (TriNetX, Inc., Cambridge, MA, USA), which provides access to de-identified electronic health record data contributed by participating healthcare institutions. Due to data use agreements, the specific datasets generated for this study cannot be downloaded or distributed outside the platform. Researchers who wish to reproduce or expand upon these analyses may obtain access to the same data through an institutional license to the TriNetX Research Network. Information regarding access can be found on the TriNetX website (https://trinetx.com/solutions/real-world-datasets/) and may vary by participating institution. Because analyses are conducted within the secure TriNetX environment using dynamically updated datasets, no static dataset, accession number, or DOI is available.

